# Making sense of the Global Coronavirus Data: The role of testing rates in understanding the pandemic and our exit strategy

**DOI:** 10.1101/2020.04.06.20054239

**Authors:** Rahul Potluri, Deepthi Lavu

**Affiliations:** ACALM Big Data Research Group

## Abstract

The Coronavirus disease 2019(COVID-19) outbreak has caused havoc across the world. Subsequently, research on COVID-19 has focused on number of cases and deaths and predicted projections have focused on these parameters. We propose that the number of tests performed is a very important denominator in understanding the COVID-19 data. We analysed the number of diagnostic tests performed in proportion to the number of cases and subsequently deaths across different countries and projected pandemic outcomes.

We obtained real time COVID-19 data from the reference website Worldometer at 0900 BST on Saturday 4^th^ April, 2020 and collated the information obtained on the top 50 countries with the highest number of COVID 19 cases. We analysed this data according to the number of tests performed as the main denominator. Country wise population level pandemic projections were extrapolated utilising three models - 1) inherent case per test and death per test rates at the time of obtaining the data (4/4/2020 0900 BST) for each country; 2) rates adjusted according to the countries who conducted at least 100000 tests and 3) rates adjusted according to South Korea.

We showed that testing rates impact on the number of cases and deaths and ultimately on future projections for the pandemic across different countries. We found that countries with the highest testing rates per population have the lowest death rates and give us an early indication of an eventual COVID-19 mortality rate. It is only by continued testing on a large scale that will enable us to know if the increasing number of patients who are seriously unwell in hospitals across the world are the tip of the iceberg or not. Accordingly, obtaining this information through a rapid increase in testing globally is the only way which will enable us to exit the COVID-19 pandemic and reduce economic and social instability.

## Introduction

The Coronavirus disease 2019(COVID-19) outbreak has caused havoc across the world after it was first reported in Wuhan, China ^1,2^. Subsequently, research on COVID-19 has exploded to understand the new disease and its impact on mankind^3-16^. However, the number of baseless articles resulting in fake news articles has also gone up exponentially^17-19^. A number of models have been adapted by policymakers to predict the course of COVID-19 across the world^4,6,13,20^. The reason for such models is to ensure that healthcare systems can plan services to help them cope with the demands of this new disease which is resulting in serious cases leading to hospitalisation^3,8^. Core elements of the prediction models have been the number of cases and deaths reported and these studies extrapolated the numbers forward to the population over time^4,6,13,20^. Given the pandemic course of COVID-19, it has become common practice to compare its spread in different countries using case fatality rates^3,4,7,13^. However, such methods only tell us part of the story. Vast differences amongst countries in their testing policies for varied reasons including availability of testing equipment, infrastructure, resources and local governing policies affect case fatality rates. In addition, comparing case fatality rates between countries which are at different stages of the epidemic in their region would be erroneous as rates at the beginning and end would be lower compared to rates at the peak when healthcare services are stretched to their limits. Therefore, the search for a common yardstick or denominator is necessary to compare different countries so that the data can be extrapolated for global comparison. Over the past four weeks, as COVID-19 spread further around the world, testing rates have picked up in most countries. We propose that analysis of the number of diagnostic tests performed in proportion to the number of cases and subsequently deaths in the underlying populations of different countries is the best way to predict what might happen next. We analysed this from the ACALM Big Data research unit.

## Methods

We obtained real time COVID-19 data from the reference website Worldometer at 0900 BST on Saturday 4^th^ April, 2020 and collated the information obtained on the top 50 countries with the highest number of COVID-19 cases^21^. From this source, we obtained many parameters including the number of country wise COVID-19 cases, deaths, tests performed, cases per million population, deaths per million population and tests per million population. China and Saudi Arabia were excluded due to lack of data on number of diagnostic tests performed, therefore numbers 51 and 52 were included in the compiled top 50 list.

We obtained case fatality rates by dividing the number of deaths by the number of cases represented as a percentage. Next, tests per positive case were calculated by dividing the number of tests by the number of cases. We then calculated the number of cases per test and number of deaths per test by dividing the number of cases and deaths respectively, by the number of tests represented as a percentage (a case per test rate and a death per test rate). Subsequently, we obtained the population of these countries (in millions) from the number of cases divided by the number of cases per million. We can obviously obtain more accurate country population statistics from other sources but to maintain our consistency of the data source and methodology (for all countries), we derived the information from this data only. We then analysed the above in three steps.

Firstly, we extrapolated the population level pandemic data for each country in terms of cases and number of deaths according to each country’s case per test rate and death per test rate as calculated as a snapshot at the time of obtaining the data.

There are a number of limitations to the methodology used when taking a snapshot of these countries at a point in time, as done above, especially because each country is likely to be on a different part of the pandemic curve and extrapolating to the population level data is not likely to be accurate. Therefore, we further undertook a consistent adjustment according to countries which performed the most tests; in favour of larger countries with bigger populations we chose an arbitrary cut off of 100,000 tests per country. 15 countries had undertaken more than 100,000 tests and as all these countries showed differences in their cases/test and deaths/test we took the 15 country group as a whole to obtain an adjustment factor according to the cases/test and deaths/test. Using this we derived a case per test rate of 13.53% and a death per test rate of 0.77% for the 15 country group. Based on the above numbers, we extrapolated figures at the population level for all 50 countries to calculate the predicated number of cases and deaths.

We felt it necessary to undertake further analysis, the third analysis, to adjust the data to a country which is progressing towards the latter half of the pandemic curve – South Korea ^22-24^. Ideally, undertaking this adjustment with data from China would be most appropriate but data for the number of diagnostic tests performed in China was not available. The adjustment factor for South Korea was a case per test rate of 2.23% and 0.04% death per test rate.

Hence our country wise population level pandemic projections were based on 1) inherent case per test and death per test rates at the time of obtaining the data (4/4/2020 0900 BST) for each country rates adjusted according to the countries who conducted at least 100000 tests and 3) rates adjusted according to South Korea. Our analyses are shown in the tables and figures. No additional analyses were performed.

## Results

Full data obtained on 4/4/2020 are shown in Table 1 for the top 50 countries with highest number of COVID-19 cases in the world. Table 2 shows the countries according to number of tests performed per positive diagnosed COVID-19 cases. Table 3 shows population level pandemic projections for cases and deaths according to each individual country’s case per test and death per test rate on 4/4/2020 0900 BST. Table 4 shows population level pandemic projections adjusted for the combined case per test and death per test rate of the 15 countries group that have performed at least 100000 tests. Table 5 shows the population level pandemic projections adjusted for the case per test and death per test rate of South Korea. Figure 1 show a scatter plot to show the relationship between the case fatality rate and the testing rate as a percentage of the total population of the country for the countries which have tested at least 1% of their total population. Italy was excluded from this scatter plot as it was an outlier with a case fatality rate of 12.25%.

**Table 1:**
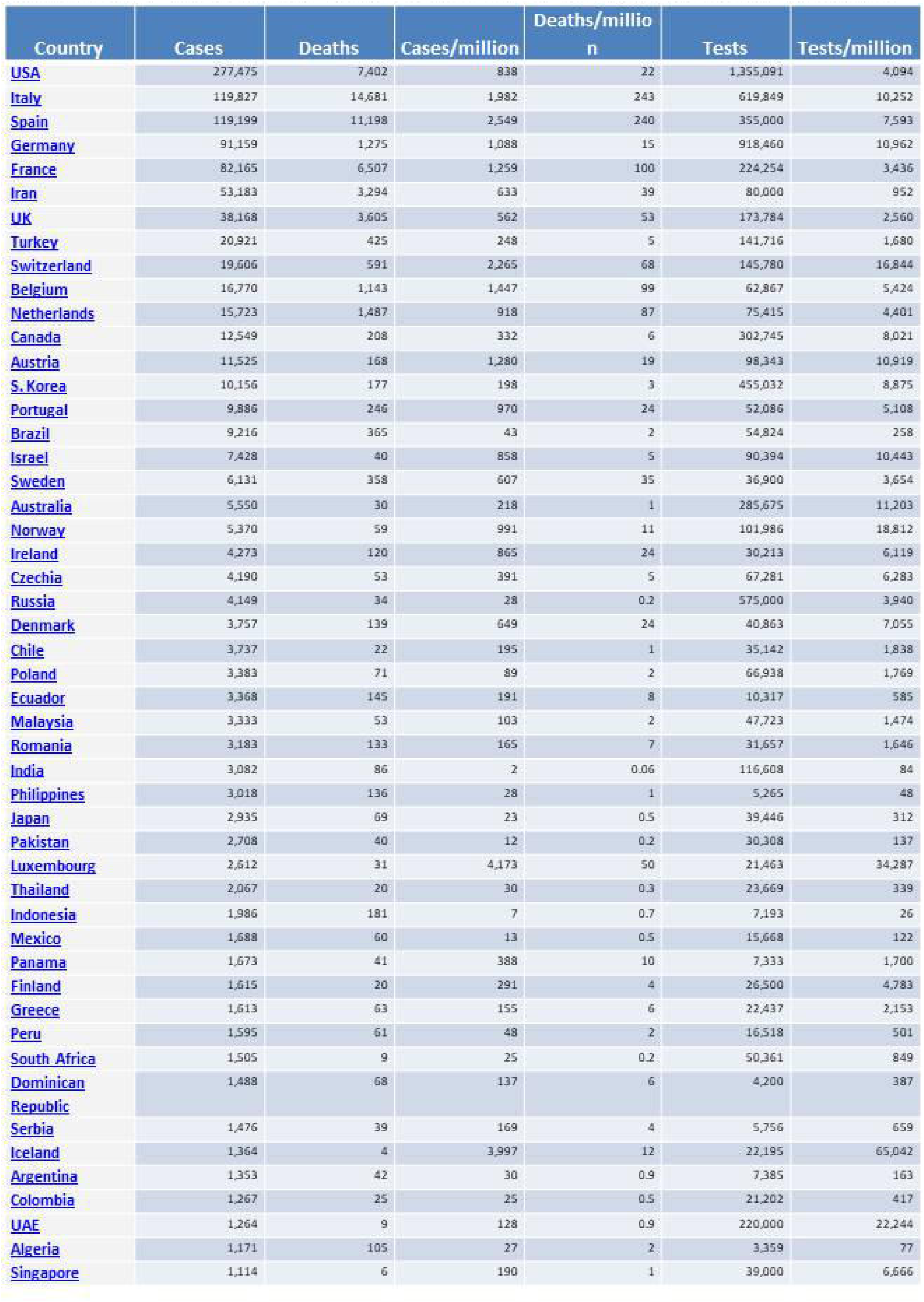
COVID-19 Data Obtained on 4/4/2020 at 0900 BST-Wordometer Coronavirus Statistics.

| Country | Cases | Deaths | Cases/million | Deaths/million | Tests | Tests/million |
| --- | --- | --- | --- | --- | --- | --- |
| USA | 277,475 | 7,402 | 838 | 22 | 1,355,091 | 4,094 |
| Italy | 119,827 | 14,681 | 1,982 | 243 | 619,849 | 10,252 |
| Spain | 119,199 | 11,198 | 2,549 | 240 | 355,000 | 7,593 |
| Germany | 91,159 | 1,275 | 1,088 | 15 | 918,460 | 10,962 |
| France | 82,165 | 6,507 | 1,259 | 100 | 224,254 | 3,436 |
| Iran | 53,183 | 3,294 | 633 | 39 | 80,000 | 952 |
| UK | 38,168 | 3,605 | 562 | 53 | 173,784 | 2,560 |
| Turkey | 20,921 | 425 | 248 | 5 | 141,716 | 1,680 |
| Switzerland | 19,606 | 591 | 2,265 | 68 | 145,780 | 16,844 |
| Belgium | 16,770 | 1,143 | 1,447 | 99 | 62,867 | 5,424 |
| Netherlands | 15,723 | 1,487 | 918 | 87 | 75,415 | 4,401 |
| Canada | 12,549 | 208 | 332 | 6 | 302,745 | 8,021 |
| Austria | 11,525 | 168 | 1,280 | 19 | 98,343 | 10,919 |
| S. Korea | 10,156 | 177 | 198 | 3 | 455,032 | 8,875 |
| Portugal | 9,886 | 246 | 970 | 24 | 52,086 | 5,108 |
| Brazil | 9,216 | 365 | 43 | 2 | 54,824 | 258 |
| Israel | 7,428 | 40 | 858 | 5 | 90,394 | 10,443 |
| Sweden | 6,131 | 358 | 607 | 35 | 36,900 | 3,654 |
| Australia | 5,550 | 30 | 218 | 1 | 285,675 | 11,203 |
| Norway | 5,370 | 59 | 991 | 11 | 101,986 | 18,812 |
| Ireland | 4,273 | 120 | 865 | 24 | 30,213 | 6,119 |
| Czechia | 4,190 | 53 | 391 | 5 | 67,281 | 6,283 |
| Russia | 4,149 | 34 | 28 | 0.2 | 575,000 | 3,940 |
| Denmark | 3,757 | 139 | 649 | 24 | 40,863 | 7,055 |
| Chile | 3,737 | 22 | 195 | 1 | 35,142 | 1,838 |
| Poland | 3,383 | 71 | 89 | 2 | 66,938 | 1,769 |
| Ecuador | 3,368 | 145 | 191 | 8 | 10,317 | 585 |
| Malaysia | 3,333 | 53 | 103 | 2 | 47,723 | 1,474 |
| Romania | 3,183 | 133 | 165 | 7 | 31,657 | 1,646 |
| India | 3,082 | 86 | 2 | 0.06 | 116,608 | 84 |
| Philippines | 3,018 | 136 | 28 | 1 | 5,265 | 48 |
| Japan | 2,935 | 69 | 23 | 0.5 | 39,446 | 312 |
| Pakistan | 2,708 | 40 | 12 | 0.2 | 30,308 | 137 |
| Luxembourg | 2,612 | 31 | 4,173 | 50 | 21,463 | 34,287 |
| Thailand | 2,067 | 20 | 30 | 0.3 | 23,669 | 339 |
| Indonesia | 1,986 | 181 | 7 | 0.7 | 7,193 | 26 |
| Mexico | 1,688 | 60 | 13 | 0.5 | 15,668 | 122 |
| Panama | 1,673 | 41 | 388 | 10 | 7,333 | 1,700 |
| Finland | 1,615 | 20 | 291 | 4 | 26,500 | 4,783 |
| Greece | 1,613 | 63 | 155 | 6 | 22,437 | 2,153 |
| Peru | 1,595 | 61 | 48 | 2 | 16,518 | 501 |
| South Africa | 1,505 | 9 | 25 | 0.2 | 50,361 | 849 |
| Dominican Republic | 1,488 | 68 | 137 | 6 | 4,200 | 387 |
| Serbia | 1,476 | 39 | 169 | 4 | 5,756 | 659 |
| Iceland | 1,364 | 4 | 3,997 | 12 | 22,195 | 65,042 |
| Argentina | 1,353 | 42 | 30 | 0.9 | 7,385 | 163 |
| Colombia | 1,267 | 25 | 25 | 0.5 | 21,202 | 417 |
| UAE | 1,264 | 9 | 128 | 0.9 | 220,000 | 22,244 |
| Algeria | 1,171 | 105 | 27 | 2 | 3,359 | 77 |
| Singapore | 1,114 | 6 | 190 | 1 | 39,000 | 6,666 |

**Table 2:**
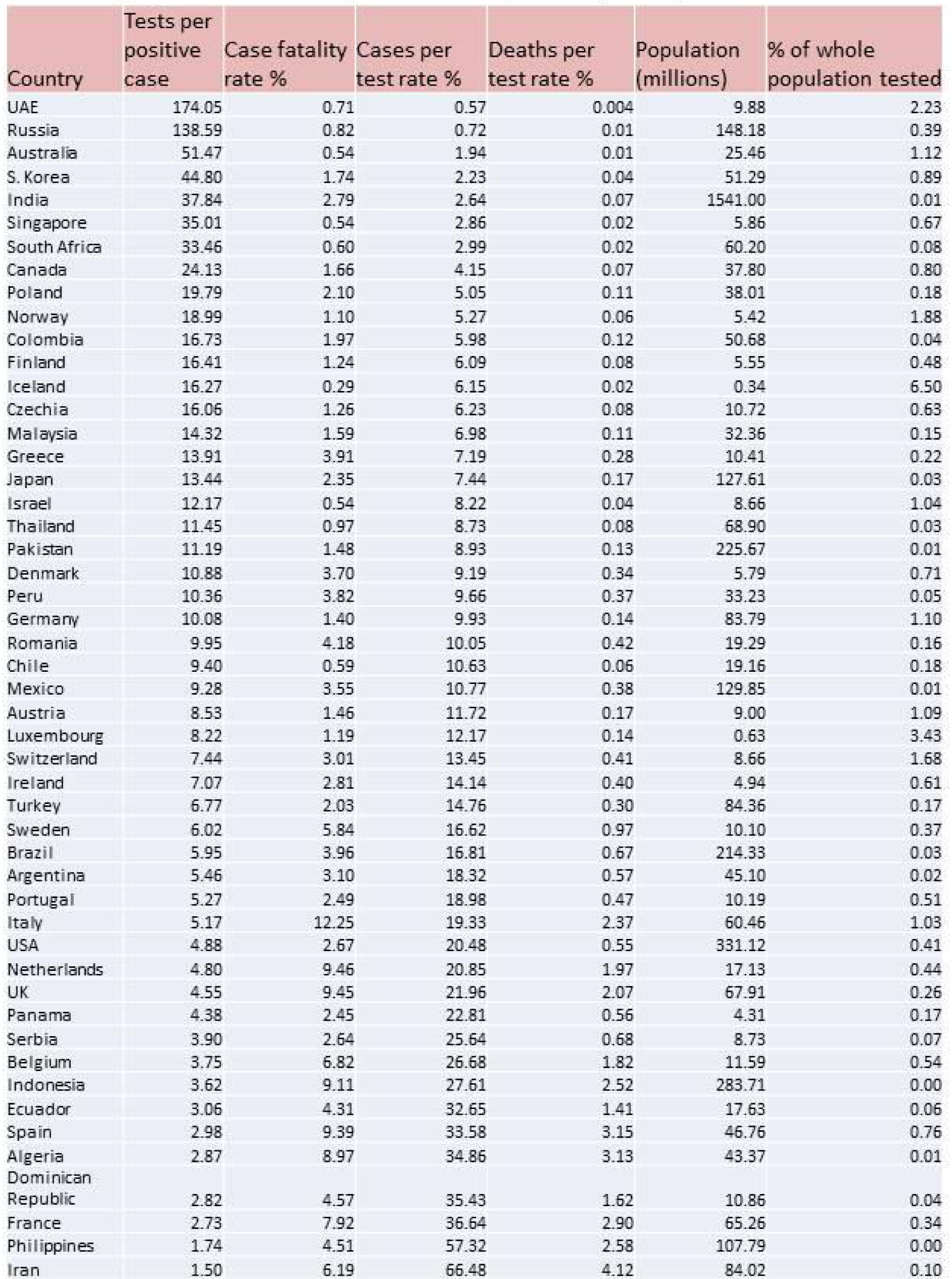
Highest COVID-19 testing rates according to tests per diagnosed case.

| Country | Tests per positive case | Case fatality rate % | Cases per test rate % | Deaths per test rate % | Population (millions) | % of whole population tested |
| --- | --- | --- | --- | --- | --- | --- |
| UAE | 174.05 | 0.71 | 0.57 | 0.004 | 9.88 | 2.23 |
| Russia | 138.59 | 0.82 | 0.72 | 0.01 | 148.18 | 0.39 |
| Australia | 51.47 | 0.54 | 1.94 | 0.01 | 25.46 | 1.12 |
| S. Korea | 44.80 | 1.74 | 2.23 | 0.04 | 51.29 | 0.89 |
| India | 37.84 | 2.79 | 2.64 | 0.07 | 1541.00 | 0.01 |
| Singapore | 35.01 | 0.54 | 2.86 | 0.02 | 5.86 | 0.67 |
| South Africa | 33.46 | 0.60 | 2.99 | 0.02 | 60.20 | 0.08 |
| Canada | 24.13 | 1.66 | 4.15 | 0.07 | 37.80 | 0.80 |
| Poland | 19.79 | 2.10 | 5.05 | 0.11 | 38.01 | 0.18 |
| Norway | 18.99 | 1.10 | 5.27 | 0.06 | 5.42 | 1.88 |
| Colombia | 16.73 | 1.97 | 5.98 | 0.12 | 50.68 | 0.04 |
| Finland | 16.41 | 1.24 | 6.09 | 0.08 | 5.55 | 0.48 |
| Iceland | 16.27 | 0.29 | 6.15 | 0.02 | 0.34 | 6.50 |
| Czechia | 16.06 | 1.26 | 6.23 | 0.08 | 10.72 | 0.63 |
| Malaysia | 14.32 | 1.59 | 6.98 | 0.11 | 32.36 | 0.15 |
| Greece | 13.91 | 3.91 | 7.19 | 0.28 | 10.41 | 0.22 |
| Japan | 13.44 | 2.35 | 7.44 | 0.17 | 127.61 | 0.03 |
| Israel | 12.17 | 0.54 | 8.22 | 0.04 | 8.66 | 1.04 |
| Thailand | 11.45 | 0.97 | 8.73 | 0.08 | 68.90 | 0.03 |
| Pakistan | 11.19 | 1.48 | 8.93 | 0.13 | 225.67 | 0.01 |
| Denmark | 10.88 | 3.70 | 9.19 | 0.34 | 5.79 | 0.71 |
| Peru | 10.36 | 3.82 | 9.66 | 0.37 | 33.23 | 0.05 |
| Germany | 10.08 | 1.40 | 9.93 | 0.14 | 83.79 | 1.10 |
| Romania | 9.95 | 4.18 | 10.05 | 0.42 | 19.29 | 0.16 |
| Chile | 9.40 | 0.59 | 10.63 | 0.06 | 19.16 | 0.18 |
| Mexico | 9.28 | 3.55 | 10.77 | 0.38 | 129.85 | 0.01 |
| Austria | 8.53 | 1.46 | 11.72 | 0.17 | 9.00 | 1.09 |
| Luxembourg | 8.22 | 1.19 | 12.17 | 0.14 | 0.63 | 3.43 |
| Switzerland | 7.44 | 3.01 | 13.45 | 0.41 | 8.66 | 1.68 |
| Ireland | 7.07 | 2.81 | 14.14 | 0.40 | 4.94 | 0.61 |
| Turkey | 6.77 | 2.03 | 14.76 | 0.30 | 84.36 | 0.17 |
| Sweden | 6.02 | 5.84 | 16.62 | 0.97 | 10.10 | 0.37 |
| Brazil | 5.95 | 3.96 | 16.81 | 0.67 | 214.33 | 0.03 |
| Argentina | 5.46 | 3.10 | 18.32 | 0.57 | 45.10 | 0.02 |
| Portugal | 5.27 | 2.49 | 18.98 | 0.47 | 10.19 | 0.51 |
| Italy | 5.17 | 12.25 | 19.33 | 2.37 | 60.46 | 1.03 |
| USA | 4.88 | 2.67 | 20.48 | 0.55 | 331.12 | 0.41 |
| Netherlands | 4.80 | 9.46 | 20.85 | 1.97 | 17.13 | 0.44 |
| UK | 4.55 | 9.45 | 21.96 | 2.07 | 67.91 | 0.26 |
| Panama | 4.38 | 2.45 | 22.81 | 0.56 | 4.31 | 0.17 |
| Serbia | 3.90 | 2.64 | 25.64 | 0.68 | 8.73 | 0.07 |
| Belgium | 3.75 | 6.82 | 26.68 | 1.82 | 11.59 | 0.54 |
| Indonesia | 3.62 | 9.11 | 27.61 | 2.52 | 283.71 | 0.00 |
| Ecuador | 3.06 | 4.31 | 32.65 | 1.41 | 17.63 | 0.06 |
| Spain | 2.98 | 9.39 | 33.58 | 3.15 | 46.76 | 0.76 |
| Algeria | 2.87 | 8.97 | 34.86 | 3.13 | 43.37 | 0.01 |
| Dominican Republic | 2.82 | 4.57 | 35.43 | 1.62 | 10.86 | 0.04 |
| France | 2.73 | 7.92 | 36.64 | 2.90 | 65.26 | 0.34 |
| Philippines | 1.74 | 4.51 | 57.32 | 2.58 | 107.79 | 0.00 |
| Iran | 1.50 | 6.19 | 66.48 | 4.12 | 84.02 | 0.10 |

**Table 3:** Highest projected population level COVID-19 deaths according to rate of death per test for each individual country on 4/4/20

| Country | Population (millions) | % of whole population tested | Extrapolated cases per population (millions) | Extrapolated deaths per population |
| --- | --- | --- | --- | --- |
| Indonesia | 283.71 | 0.003 | 78.33 | 7139203 |
| Iran | 84.02 | 0.10 | 55.85 | 3459416 |
| Philippines | 107.79 | 0.00 | 61.78 | 2784208 |
| France | 65.26 | 0.34 | 23.91 | 1893659 |
| USA | 331.12 | 0.41 | 67.80 | 1808675 |
| Spain | 46.76 | 0.76 | 15.70 | 1475078 |
| Italy | 60.46 | 1.03 | 11.69 | 1431927 |
| Brazil | 214.33 | 0.03 | 36.03 | 1426909 |
| UK | 67.91 | 0.26 | 14.92 | 1408830 |
| Algeria | 43.37 | 0.01 | 15.12 | 1355728 |
| India | 1541.00 | 0.01 | 40.73 | 1136509 |
| Mexico | 129.85 | 0.01 | 13.99 | 497241 |
| Netherlands | 17.13 | 0.44 | 3.57 | 337712 |
| Pakistan | 225.67 | 0.01 | 20.16 | 297831 |
| Argentina | 45.10 | 0.02 | 8.26 | 256493 |
| Turkey | 84.36 | 0.17 | 12.45 | 252989 |
| Ecuador | 17.63 | 0.06 | 5.76 | 247830 |
| Japan | 127.61 | 0.03 | 9.49 | 223217 |
| Belgium | 11.59 | 0.54 | 3.09 | 210711 |
| Dominican Republic | 10.86 | 0.04 | 3.85 | 175850 |
| Peru | 33.23 | 0.05 | 3.21 | 122713 |
| Germany | 83.79 | 1.10 | 8.32 | 116311 |
| Sweden | 10.10 | 0.37 | 1.68 | 97994 |
| Romania | 19.29 | 0.16 | 1.94 | 81047 |
| Colombia | 50.68 | 0.04 | 3.03 | 59759 |
| Serbia | 8.73 | 0.07 | 2.24 | 59176 |
| Thailand | 68.90 | 0.03 | 6.02 | 58220 |
| Portugal | 10.19 | 0.51 | 1.93 | 48135 |
| Poland | 38.01 | 0.18 | 1.92 | 40318 |
| Malaysia | 32.36 | 0.15 | 2.26 | 35937 |
| Switzerland | 8.66 | 1.68 | 1.16 | 35092 |
| Greece | 10.41 | 0.22 | 0.75 | 29220 |
| Canada | 37.80 | 0.80 | 1.57 | 25969 |
| Panama | 4.31 | 0.17 | 0.98 | 24108 |
| S. Korea | 51.29 | 0.89 | 1.14 | 19952 |
| Denmark | 5.79 | 0.71 | 0.53 | 19692 |
| Ireland | 4.94 | 0.61 | 0.70 | 19620 |
| Austria | 9.00 | 1.09 | 1.06 | 15381 |
| Chile | 19.16 | 0.18 | 2.04 | 11997 |
| South Africa | 60.20 | 0.08 | 1.80 | 10758 |
| Russia | 148.18 | 0.39 | 1.07 | 8762 |
| Czechia | 10.72 | 0.63 | 0.67 | 8442 |
| Finland | 5.55 | 0.48 | 0.34 | 4189 |
| Israel | 8.66 | 1.04 | 0.71 | 3831 |
| Norway | 5.42 | 1.88 | 0.29 | 3135 |
| Australia | 25.46 | 1.12 | 0.49 | 2674 |
| Luxembourg | 0.63 | 3.43 | 0.08 | 904 |
| Singapore | 5.86 | 0.67 | 0.17 | 902 |
| UAE | 9.88 | 2.23 | 0.06 | 404 |
| Iceland | 0.34 | 6.50 | 0.02 | 62 |

**Table 4:**
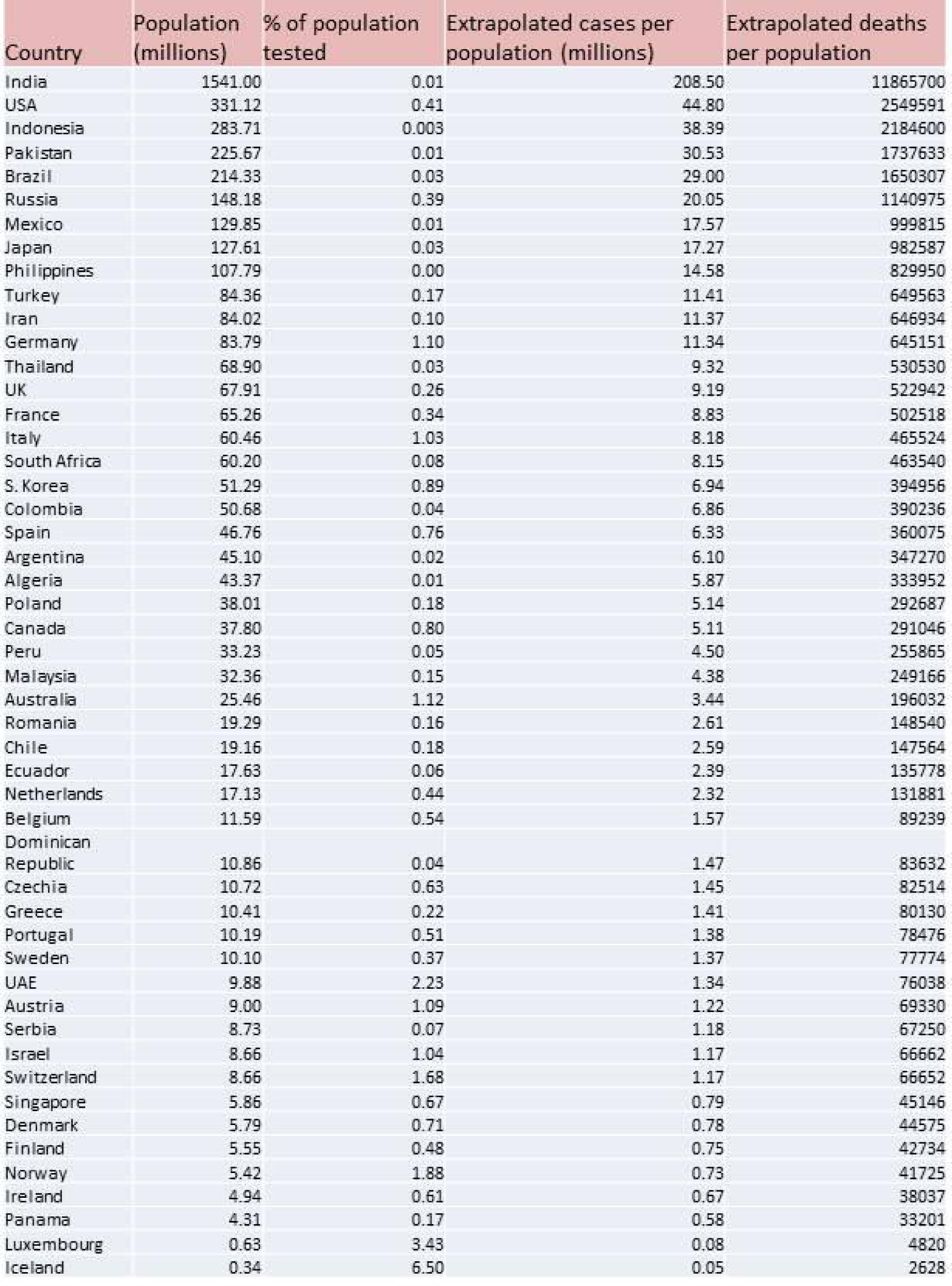
Highest projected population level COVID-19 deaths after adjustment according to countries who have tested at least 100000 tests

**Table 5:** Highest projected population level COVID-19 deaths after adjustment according to South Korea.

| Country | Population (millions) | % of population tested | Extrapolated cases per population (millions) | Extrapolated deaths per population |
| --- | --- | --- | --- | --- |
| India | 1541.00 | 0.01 | 34.36 | 616400 |
| USA | 331.12 | 0.41 | 7.38 | 132446 |
| Indonesia | 283.71 | 0.003 | 6.33 | 113486 |
| Pakistan | 225.67 | 0.01 | 5.03 | 90267 |
| Brazil | 214.33 | 0.03 | 4.78 | 85730 |
| Russia | 148.18 | 0.39 | 3.30 | 59271 |
| Mexico | 129.85 | 0.01 | 2.90 | 51938 |
| Japan | 127.61 | 0.03 | 2.85 | 51043 |
| Philippines | 107.79 | 0.00 | 2.40 | 43114 |
| Turkey | 84.36 | 0.17 | 1.88 | 33744 |
| Iran | 84.02 | 0.10 | 1.87 | 33607 |
| Germany | 83.79 | 1.10 | 1.87 | 33514 |
| Thailand | 68.90 | 0.03 | 1.54 | 27560 |
| UK | 67.91 | 0.26 | 1.51 | 27166 |
| France | 65.26 | 0.34 | 1.46 | 26105 |
| Italy | 60.46 | 1.03 | 1.35 | 24183 |
| South Africa | 60.20 | 0.08 | 1.34 | 24080 |
| S. Korea | 51.29 | 0.89 | 1.14 | 19952 |
| Colombia | 50.68 | 0.04 | 1.13 | 20272 |
| Spain | 46.76 | 0.76 | 1.04 | 18705 |
| Argentina | 45.10 | 0.02 | 1.01 | 18040 |
| Algeria | 43.37 | 0.01 | 0.97 | 17348 |
| Poland | 38.01 | 0.18 | 0.85 | 15204 |
| Canada | 37.80 | 0.80 | 0.84 | 15119 |
| Peru | 33.23 | 0.05 | 0.74 | 13292 |
| Malaysia | 32.36 | 0.15 | 0.72 | 12944 |
| Australia | 25.46 | 1.12 | 0.57 | 10183 |
| Romania | 19.29 | 0.16 | 0.43 | 7716 |
| Chile | 19.16 | 0.18 | 0.43 | 7666 |
| Ecuador | 17.63 | 0.06 | 0.39 | 7053 |
| Netherlands | 17.13 | 0.44 | 0.38 | 6851 |
| Belgium | 11.59 | 0.54 | 0.26 | 4636 |
| Dominican Republic | 10.86 | 0.04 | 0.24 | 4345 |
| Czechia | 10.72 | 0.63 | 0.24 | 4286 |
| Greece | 10.41 | 0.22 | 0.23 | 4163 |
| Portugal | 10.19 | 0.51 | 0.23 | 4077 |
| Sweden | 10.10 | 0.37 | 0.23 | 4040 |
| UAE | 9.88 | 2.23 | 0.22 | 3950 |
| Austria | 9.00 | 1.09 | 0.20 | 3602 |
| Serbia | 8.73 | 0.07 | 0.19 | 3493 |
| Israel | 8.66 | 1.04 | 0.19 | 3463 |
| Switzerland | 8.66 | 1.68 | 0.19 | 3462 |
| Singapore | 5.86 | 0.67 | 0.13 | 2345 |
| Denmark | 5.79 | 0.71 | 0.13 | 2316 |
| Finland | 5.55 | 0.48 | 0.12 | 2220 |
| Norway | 5.42 | 1.88 | 0.12 | 2168 |
| Ireland | 4.94 | 0.61 | 0.11 | 1976 |
| Panama | 4.31 | 0.17 | 0.10 | 1725 |
| Luxembourg | 0.63 | 3.43 | 0.01 | 250 |
| Iceland | 0.34 | 6.50 | 0.01 | 137 |

**Figure 1:**
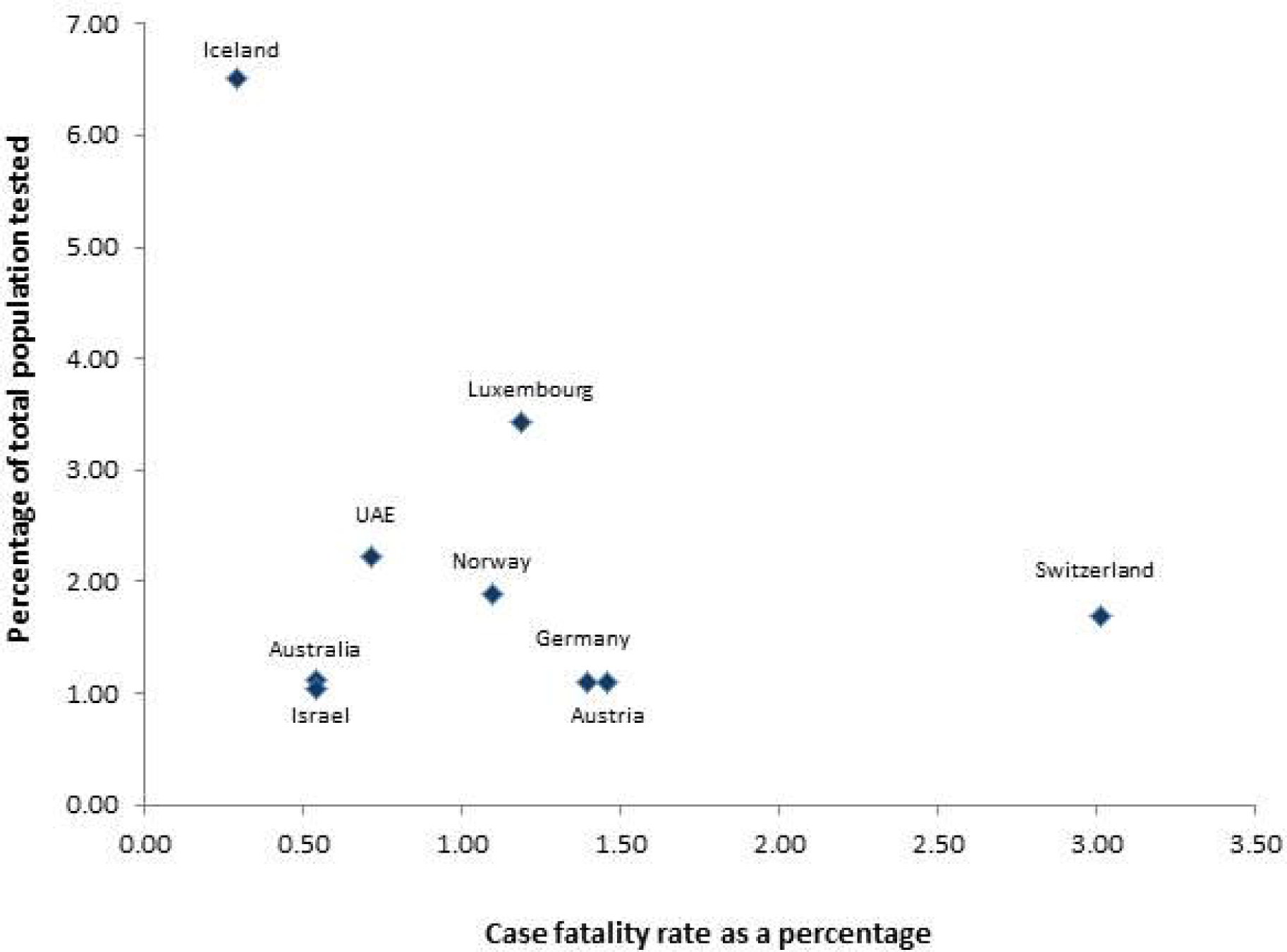
Case fatality rates according to percentage of total population tested for countries who have tested at least 1% of the whole population excluding Italy

## Discussion

COVID-19 statistics are complex and comparing different countries based on number of total cases, deaths and/or case fatality rate does not show the complete picture (Table 1). A common denominator is required to make senses of these numbers and we propose that this denominator is the number of diagnostic tests performed. In our analyses we showed the deaths and cases in relation to the number of tests performed and presented population level pandemic projections based on these. This is particularly relevant in the current environment where testing parameters vary across different countries leading to non-uniformity in projections. It is important to discuss each of our different analyses in turn, the rationale, drawbacks and what it means for different countries.

As table 2 shows, the number of tests per positive case is an important parameter because it is an indication of how widely the testing policy of the respective country has followed the advice from the World Health Organisation (WHO)^1^. Analysis using tests per positive case approach favours richer countries with smaller populations such as the UAE which tests over 174 people per positive case. However, there are exceptions to this such as Russia and India which both have large populations. This data suggests that both countries are undertaking a large number of tests to detect one positive case. In these countries the overall percentage of population tested is low. Bias in these figures could be the reliability of the reported number of tests performed. For example, the figures for India as released by the Indian Council of Medical Research^25^ in terms of numbers tested are not as high as the raw data obtain from this source but for consistency in dealing with all the raw data in the same way, we analysed according to the data obtained from Worldometer. Of course similar bias could be inherent for the testing data for all countries but in our defence we have treated all the raw data obtained in the same way for consistency and have opened up the data for scrutiny. Another important factor to consider here is the testing policy followed in these countries. Are the countries at the top of this table testing a cohort of people who have a low possibility of carrying this infection? If only those with symptoms are tested then individuals are more likely to test positive for COVID-19 leading to a low test per positive number. If these countries test the sickest of patients as you would expect in countries with the largest populations and limited testing kits to do, then high testing rates per positive case is even more remarkable as it may suggest lower virus rates compared to other countries but this cannot be concluded from this study. Furthermore, the reliability of local testing kits is an important factor as there are a number of reports of COVID-19 patients testing negative numerous times before a positive test^11^. South Korea can be considered as an exception to this because following an explosion of cases initially, they embarked on an extensive testing policy along with isolation policies combined with the utility of mobile tech and applications to inform the public about real time locations of positive cases. As such, it is widely accepted that South Korea are further along the pandemic curve and the rates of new cases and deaths have significantly reduced ^22-24^.

Projections for the pandemic on an individual population level are very important for governments to plan and organise healthcare systems in response. COVID-19 presents a unique problem because there is no immunity for this in the community, nor a vaccination or targeted medical treatment. Given that this is a highly contagious disease that spreads very quickly, if a large part of the population suffer from the disease in a short space of time, even if majority of cases are mild, a small minority of severe/critical cases will still lead to significant pressures on healthcare systems as now seen in Italy, Spain and the USA (particularly New York). Globally lockdowns have been instated to reduce the spread of infection, allow the healthcare systems to cope with the condition and “flatten the curve” of the pandemic. These were not enforced all at once and the projections in table 3 are based on the data available on 4/4/2020 and a snapshot depending on the actions, policies of individual countries. Much more complex models have been undertaken by different groups which included time as a variable ^20^. However, we propose that testing rate is a very important parameter in projecting the outcomes of the pandemic. Therefore, whilst it seems far-fetched to suggest that Indonesia may end up with over 7 million deaths from less than 2000 cases reported so far, we have to note that only just over 7000 tests have been performed for a country of 283 million. There are a number of factors for low testing rates such as local policies, lack of resources and equipment and it is impossible to discuss them all; we point out that testing rates are extremely important in projecting the outcomes of the pandemic particularly in countries with large populations. In this context, if we look at countries such as the UK and India both of which have tested over 100000 tests, given the large populations and their case per test rate and death per test rate on 4/4/2020, both countries have projections for over 1000000 deaths.

As mentioned the position of any given country on the pandemic curve is important in determining population level projections and since we proposed that testing rates have an impact on projections we adjusted all projections to the combined case per test and death per test rates of all the countries that have performed over 100000 tests. This analyses is shown in table 4. We also felt that projections should be done on the case per test and death per test rate for South Korea given the countries position on the pandemic curve and are shown in table 4^22-24^. Both these analyses are biased in terms of predications for total deaths for countries with larger populations. For example in spite of the case per test and deaths per test rate being low in India, as the population of the country is large, the projections are still over 10 millions deaths as per table 4 (adjustment according to countries which have performed more than 100,000 tests) and 500,000 deaths as per table 5 (South Korea adjustment).

It is also no coincidence that none of the top 10 countries in table 4 or table 5 have tested at least 1% of the total population. We looked at the countries that have tested at least 1% of their populations and looked at their cases fatality rate. We excluded the 10^th^ country on the list – Italy because of its high case fatality rate of 12.25%. All other countries had a case fatality rate of 3% or lower. We then correlated the case fatality rate with percentage of the population tested as shown in figure 1. This approach showed higher percentage of population tested in countries with lower populations who have tested a higher proportion of their total population, but not in all cases. Both Germany (population 83 million) and to a lesser extent Australia (population 25million) and have tested more than 1% of their population and showed low case fatality rates (1.4% Germany and 0.54% Australia). Provided that their testing criteria is reliable, these figures may serve as early indicators of the actual mortality rate for COVID-19 and these low figures are encouraging.

None of these methods used for projections are likely to hold true in reality. If we go back to the analyses for South Korea and its projections of approximately 20,000 deaths, there have been only 177 deaths in South Korea so far. It seems highly improbable that for a country where the number of cases and deaths have significantly tailed off would end up with 19,952 deaths. Furthermore, the herd immunity concept has been a strategy to contain disease spread not only for COVID-19 but across a number of pandemics such as Swine Flu ^26^. Although it is widely debated as to what percentage of the population would need to be affected by the disease to confer herd immunity, a figure of 60% has been widely used^27-29^. Even if we adjust the South Korea figures (table 5) to 60%, we will probably still over estimate the number of deaths.

Where does all of this leave us and what is the point of all these statistics and analyses? Clearly from the example of South Korea we can contain COVID-19 and in spite of differences of the specific policies of lockdown between countries, social distancing and limiting spread are the broad themes to take forward. The analyses in this study highlight the importance of testing as the relevant denominator for which all the COVID-19 data should be related to. The testing policy is advocated strongly by the WHO in their COVID-19 statements^1^. The suggested early indication of a low mortality rate from our analyses, coupled with the fact that COVID-19 is a new disease affecting the globe in a short time, it is highly plausible that the serious cases and deaths we are seeing in the some countries may be the tip of the iceberg of a disease that has spread widely. If we look at influenza data there are millions of cases and up to half a million deaths worldwide every year due to flu and these tend to be seasonal in spite of vaccination programmes and herd immunity to some extent ^27-30^. In the case of COVID-19 we might be experiencing the full whammy of a disease without immunity, globally all at once resulting in deaths. The magnitude of these deaths in perspective to other diseases such as Influenza may not be high^30^. Our analyses in this study do not prove this theory but the only thing that can is continued extensive and rapid testing across the globe. This may be the only exit strategy to prevent COVID-19 related economic and social breakdown.

## Data Availability

All data obtained and used for analysis is shown in the tables for scrutiny.

